# Dalbavancin in real life: Economic impact of prescription timing in French hospitals

**DOI:** 10.1101/2021.07.24.21259935

**Authors:** Guillaume Béraud, Jean-Claude Maupetit, Audric Darras, Alexandre Vimont, Martin Blachier

## Abstract

**Objectives:** The extended half-life of dalbavancin justifies a once-a-week dosing schedule and is supposed to favour early discharge. These advantages may therefore compensate for the cost of dalbavancin. We aimed to assess the real-life budget impact of dalbavancin through its impact on the length of stay in French hospitals.

**Methods:** A multicentre cohort based on the French registry of dalbavancin use in 2019 was compared to the French national discharge summary database. Lengths of stay and budget impact related to the infection type, the time of introduction of dalbavancin, the type of catheter and patient subgroups were assessed. An early switch was defined when dalbavancin was administered as the first or second treatment and within less than 11 days of hospitalization.

**Results:** One hundred seventy-nine patients were identified in the registry, and 154 were included in our study. Dalbavancin is mostly used for bone and joint infections, infective endocarditis and acute bacterial skin and skin structure infections. When compared to the data for similar patients in the national database, the length of stay was almost always shorter for patients treated with dalbavancin. The budget impact for dalbavancin was heterogeneous but frequently generated savings. Early switching was associated with savings (or lesser costs). Patients who required a deep venous catheter and those with the most severe patients benefited the most from dalbavancin.

**Conclusions:** Our study confirms that dalbavancin is associated with early discharge, which can offset its cost and generate savings. The greatest benefit is achieved with an early switch.

## Introduction

Antimicrobial resistance among gram-positive bacteria, particularly staphylococci and enterococci, results in considerable morbidity, mortality and cost. However, resistance to dalbavancin is rare, its efficacy is noninferior to other anti-gram-positive antibiotics, and tolerance is usually better [1]. Furthermore, the extended half-life of dalbavancin of 14 days, compared to less than 12 h for vancomycin and daptomycin, is advantageous and justifies a once-a-week dosing schedule (or even more spaced) when more than 1 injection is required. Therefore, dalbavancin offers a quality-of-life gain for patients, eliminates the need for a central catheter and the risk of potential associated complications, increases compliance and reduces the need for drug monitoring. This prolonged half-life is also supposed to favour early discharge for patients requiring parenteral treatment. These advantages may therefore compensate for the cost of dalbavancin. Indeed, Keyloun et al theorized and modelled the economic benefit of long-acting antibiotics for acute bacterial skin and skin structure infections (ABSSSIs) in an emergency department [2]. In addition, as long-acting antibiotics seem ideal for chronic infections requiring prolonged antibiotic treatment, dalbavancin has been used successfully for off-label indications such as bone and joint infections (BJIs) [3], for complex situations in which the pathogen is resistant to oral antibiotics or the predictable adherence is low, and more recently for infective endocarditis (IE) [4,5]. In fact, the only randomized controlled trial (RCT) comparing dalbavancin to standard of care for BJIs confirmed efficacy, good tolerance and a significantly shorter length of stay (LoS) [6]. While the tolerance, efficacy and effect on LoS of dalbavancin have been largely confirmed for on- and off-label indications [4–11], the associated economic benefit is mostly based on simulations [12–14]. Most economic studies have extrapolated the reduction in the LoS observed in real-life studies compared to usual treatments to a reduction in hospitalization-related costs from a societal perspective [15–17]. A budget impact analysis of dalbavancin use in a small series of twelve self-pay patients treated for ABSSSIs [18] was carried out, but its specific context (self-pay patients in the USA) made it hardly extrapolatable. To date, no comparison between dalbavancin and standard of care in real patients has been carried out with an economic analysis on the impact on the LoS. Real-life use and budget impact are poorly described; therefore, we aimed to assess the use of dalbavancin in French hospitals and to estimate the resulting average LoS and budget impact. We also expected to provide recommendations for the optimal use of dalbavancin from a health economic perspective.

## Materials and methods

We conducted an observational, retrospective, multicentre study describing the use of dalbavancin in France. This cohort was based on the French registry of dalbavancin (under the initiative of UniHA, promoted by CORREVIO) from 2019. UniHA is the largest buyer’s cooperative network for French hospitals, with more than 900 French hospitals and 96 territorial hospital groups. Twenty-four hospitals from metropolitan France, among which twelve university hospitals (supplementary figure 1), reported patients who received dalbavancin between January 1, 2019, and December 31, 2019. Factors associated with the LoS were assessed through univariate and multivariate analyses with a stepwise selection of variables. This cohort was then compared to the French National Hospital discharge summary database (*Programme de médicalisation des systèmes d’information: PMSI*) [19] and stratified according to the patient subgroup and type of venous access.

Multiple situations were explored based on the following:

- Infection type: BJI, IE or ABSSSI.
- Time before the first injection of dalbavancin: ≤ 7 days, ≤ 11 days, ≤ 25 days and any timing.
- Dalbavancin as a first or second line of treatment or any line of treatment.
- Availability and type of catheter: Deep venous catheter (implanted port), transcutaneous catheter (peripherally inserted central catheter) or general population regardless of the presence of a catheter.
- Patient subgroups: Diagnosis-related group (DRGs) defining a global severity of patient conditions (1: none, 2: mild, 3: moderate, 4: severe).

DRGs are classifications of hospitalized patients according to diseases and patient comorbidities, which provides a basis for calculating a hospital’s reimbursement to cover hospitalization cost [20]. If the LoS of a patient is shorter than the average, the fee is relatively favourable, but as the LoS increases beyond the average LoS for a specific DRG, then the cost becomes increasingly less favourable. Budget impact analysis was carried out with a microcosting approach, hospital perspective and LoS horizon. The LoS of each DRG patient from the registry was compared to the LoS of patients with a similar DRG in the national database. The revenue based on the DRG was assessed according to the global cost associated with the specific DRG (which takes into account the severity of the patient’s condition and the presence and type of catheter, among other things), calculated per hospitalization day, which consequently reflects increases in LoS. The budget impact was calculated by retrieving the additional cost of using dalbavancin instead of its comparators - vancomycin or daptomycin - and the revenue generated by the patient according to the DRG.

We defined the early switch when dalbavancin was administered as the first- or second-line treatment and within less than 7 days or 11 days of hospitalization. Comparison according to the type of catheter was achieved by comparing every patient from our cohort to a group of patients with similar DRGs in the national database who benefitted from a specific catheter according to the Classification Commune des Actes Médicaux (CCAM) (medical classification for clinical procedures) (deep venous catheter (port or port-a-cath) (CCAM code: EBLA003), transcutaneous catheter (PICC line) (CCAM code: EPLF002) or to the whole group of patients with a similar DRG regardless of the catheter (general population)). Comparisons according to global patient severity were carried out by focusing on DRGs with a severity of 3 (moderate) or 4 (severe).

Additional costs related to vancomycin administration (infusion system, therapeutic drug monitoring, side effects, etc.) were not taken into account. The cost per vial and daily cost were 3.6 € and 7.3 € for 1 g of vancomycin (prescribed at 2 g/day), 45.0 € for daptomycin (8 mg/kg/d for an average weight of 70 kg) and 2211.6 € and 4423.2 € for dalbavancin (3 doses of 500 mg, 2 injections on the first day of treatment), respectively.

Quantitative variables are described as the median (min-max) when not mentioned otherwise. LoS are reported as the median (Q1-Q3) because of a skewed distribution. Analyses were carried out with SAS (version 9.4). The study was approved by the French data protection board (CNIL MR004: n°2213417).

## Results

The UNIHA cohort included 179 patients treated with dalbavancin in 2019; 154 of these patients were included in the analysis. Twenty-five patients were excluded (11 were outpatients, 6 started their treatment before hospitalization, 5 were hospitalized in 2018, and 3 were treated after hospitalization). Overall, 105 (68%) patients were men aged 69.0 (15-94) years, with a body mass index (BMI) of 25.9 (14.3-56.4) kg/m^2^ and 3 (0-40) comorbidities, most frequently cardiovascular.

Dalbavancin was mostly used for BJIs (56%), IE (19%) and ABSSSIs (6%) (Table 1) at 3000 (1000; >4500) mg. The most frequent pathogens were methicillin-resistant *Staphylococcus epidermidis* (33%), methicillin-sensitive *S. aureus* (25%) and methicillin-resistant *S. aureus* (15%) (Supplementary Table 1). Patients received 1 (0-10) antimicrobial treatment before dalbavancin 11 (0-100) days after hospitalization, which resulted in an LoS of 18 (2-311) days (1^st^; 3^rd^ quartiles: 8.0; 34.0 days) (Table 1).

**Table 1:** Characteristics of patients according to the site of infection, treatment strategy and length of stay. Quantitative variables are expressed as the median (min-max).

|  | n (%) | Male (%) | BMI | Age | Number of comorbidities | Median (min-max) number of treatments before dalbavancin | Dalbavancin dose | Timing between hospitalization and injection | Timing between injection and end of stay | Length of stay |
| --- | --- | --- | --- | --- | --- | --- | --- | --- | --- | --- |
| Bloodstream infection | 10 (6) | 8 (80) | 25.5 (18.9-34.3) | 65.3 (28-92) | 6.1 (1-31) | 1.5 (0-10) | 1500 (1500 - 3000) | 12 (4-33) | 2.5 (0-21) | 20 (4-43) |
| Infective endocarditis | 29 (19) | 23 (79) | 24.4 (16.8-43.0) | 66.9 (21-91) | 5.1 (1-36) | 2 (0-3) | 2000 (1000 - >4500) | 13 (0-71) | 6 (0-49) | 22 (3-96) |
| Bone and joint infections | 86 (56) | 59 (69) | 26.1 (14.3-45.2) | 63.3 (15-94) | 4.5 (0-40) | 1 (0-3) | 3000 (1000 - >4500) | 11 (0-100) | 4 (0-69) | 19 (2-105) |
| Soft tissue infections | 10 (6) | 4 (40) | 25.0 (19.5-56.4) | 60.7(24-80) | 7.2 (0-24) | 1 (0-6) | 2500 (1500 - >4500) | 6 (0-34) | 7 (0-277) | 11 (2-311) |
| Catheter-related bloodstream infections | 9 (6) | 3 (33) | 26.6 (18.8-35.6) | 64.3 (25-83) | 4.1 (1-18) | 1 (0-2) | 1500 (1500-1500) | 6 (0-34) | 5 (1-11) | 9 (3-43) |
| Endovascular infections | 2 (1) | 2 (100) | 23.7 (21.8-25.6) | 70 (69-71) | 1.5 (1-2) | 2.5 (0-3) | 2250 (1500-3000) | 15 (4-26) | 1.5 (1-2) | 16.5 (5-28) |
| Others | 6 (4) | 1 (50) | 32.5 (22.2-39.0) | 59.7 (21-78) | 2.5 (1-4) | 2.5 (2-3) | 2500 (1500-3000) | 18 (1-97) | 11.5 (1-34) | 35.5 (2-107) |
| Missing | 2 (1) | 5 (83) | 28.3 (19.0-37.7) | 54 (35-73) | 4.5 (3-6) | 1(1-1) | 2250 (1500->4500) | 16.5 (13-20) | 21 (0-42) | 37.5 (13-62) |
| Total | 154 (100) | 105 (68) | 25.5 (14.3-56.4) | 63.8 (15-94) | 4.7 (0-40) | 1(0-10) | 3000 (1000->4500) | 11 (0-100) | 4 (0-277) | 18 (2-311) |

Among the 125 patients treated for the 3 main indications (BJIs, IE and ABSSSIs), 13 (10%) and 56 (45%) received dalbavancin as a first- and second-line treatment, respectively, and 12 and 39 patients specifically received dalbavancin for BJIs. Five antimicrobials, namely, daptomycin, amoxicillin, linezolid, vancomycin and teicoplanin, alone or in combination, represented 77% of the anti-gram-positive cocci antibiotics used as a first-line treatment before dalbavancin for the 3 main indications and 55% when used specifically for BJIs (Supplementary Table 2). No factors were significantly associated with the LoS in univariate analysis, except for *Enterococcus faecalis*, which was associated with a longer LoS (Table 2). The multivariate analysis suggested that the LoS increased with age, the number of treatments, the number of comorbidities, obesity, and infection due to *E. faecalis* but also indicated numerous interactions, showing the extreme heterogeneity of the patients treated with dalbavancin. As an example, the patient with the longest LoS (311 days) had no risk factors (Table 1 and Supplementary Table 1). Taking into account all the interactions would require splitting the cohort into a half-dozen subgroups and would make the multivariate analysis results unreliable.

**Table 2.** Univariate analysis. For all patients (n=154), the median [Q1-Q3] length of stay was 18.0 [8.0; 34.0] days.

| Variable | Length of stay (median [Q1-Q3]) |  |  |
| --- | --- | --- | --- |
|  | Presence of the factor | Absence of the factor | p-value |
| Sex (Male) | 17.0 [8.0; 34.0] (n=105) | 21.0 [9.0; 43.0] (n=49) | 0.10 |
| Age: > 60y | 18.0 [8.5; 39.5] (n=104) | 17.0 [8.0; 33.0] (n=50) | 0.53 |
| > 1 comorbidity | 17.5 [8.0; 30.0] (n=50) | 18.0 [9.0; 36.0] (n=104) | 0.63 |
| Diabetes | 13.0 [7.5; 28.5] (n=24) | 19.0 [9.0; 41.0] (n=130) | 0.14 |
| Obesity | 23.0 [11.0; 46.0] (n=49) | 16.0 [7.0; 33.0] (n=105) | 0.09 |
| IV drug user | 22.5 [13.0; 33.0] (n=10) | 18.0 [8.0; 36.0] (n=144) | 0.77 |
| Cancer | 16.0 [7.0; 28.0] (n=23) | 19.0 [9.0; 39.0] (n=131) | 0.63 |
| Cirrhosis | 26.0 [7.0; 33.0] (n=6) | 18.0 [8.5; 36.0] (n=148) | 0.83 |
| Polyarthritis | 24.0 [19.0; 34.0] (n=13) | 17.0 [8.0; 34.0] (n=141) | 0.62 |
| Immunosuppression | 23.0 [7.0; 34.0] (n=10) | 18.0 [8.5; 36.0] (n=144) | 0.68 |
| Hypertension | 24.0 [8.0; 43.0] (n=58) | 17.0 [8.5; 31.5] (n=96) | 0.73 |
| Other comorbidity | 21.5 [9.0; 43.0] (n=76) | 15.0 [8.0; 29.0] (n=78) | 0.28 |
| > 1 treatment before dalbavancin | 16.0 [8.0; 34.0] (n=85) | 21.0 [10.0; 38.0] (n=69) | 0.91 |
| Bloodstream infection | 20.0 [9.0; 24.0] (n=10) | 18.0 [8.0; 38.5] (n=144) | 0.47 |
| Infective endocarditis | 22.0 [13.0; 42.0] (n=29) | 18.0 [8.0; 33.0] (n=125) | 0.76 |
| Bone and joint infection | 19.0 [9.0; 34.0] (n=86) | 16.0 [7.0; 41.5] (n=68) | 0.49 |
| Soft tissues infection | 11.0 [6.0; 21.0] (n=10) | 18.5 [9.0; 36.0] (n=144) | 0.20 |
| Catheter-related bloodstream infection | 9.0 [7.0; 14.0] (n=9) | 19.0 [9.0; 38.0] (n=145) | 0.22 |
| Endovascular infection | 16.5 [5.0; 28.0] (n=2) | 18.0 [8.5; 36.0] (n=152) | 0.64 |
| Other | 35.5 [10.0; 80.0] (n=8) | 18.0 [8.0; 34.0] (n=146) | 0.11 |
| SASM | 17.0 [9.0; 41.0] (n=35) | 18.0 [8.0; 34.0] (n=119) | 0.97 |
| SARM | 24.0 [9.0; 42.0] (n=23) | 17.0 [8.0; 34.0] (n=131) | 0.23 |
| SCN | 15.0 [12.0; 28.0] (n=15) | 18.0 [8.0; 39.0] (n=139) | 0.45 |
| SERM | 21.0 [9.0; 33.0] (n=51) | 16.0 [8.0; 39.0] (n=103) | 0.92 |
| Streptococcus spp. | 20.0 [12.0; 34.0] (n=13) | 18.0 [8.0; 34.0] (n=141) | 0.88 |
| E. faecium | 43.0 [8.0; 52.0] (n=7) | 18.0 [8.0; 34.0] (n=147) | 0.61 |
| E. faecalis | 34.0 [11.0; 64.0] (n=17) | 17.0 [8.0; 33.0] (n=137) | 0.02 |
| Corynebacterium acnes | 11.0 [9.0; 31.0] (n=13) | 18.0 [8.0; 38.0] (n=141) | 0.33 |
| Undocumented infection | 11.0 [5.0; 18.0] (n=6) | 18.5 [8.5; 36.0] (n=148) | 0.34 |
| Other pathogen | 31.5 [6.0; 52.0] (n=15) | 17.5 [9.0; 33.5] (n=139) | 0.52 |
| Polymicrobial | 16.5 [8.0; 33.0] (n=116) | 25.0 [9.0; 46.0] (n=38) | 0.32 |

In addition, thirty-six participants for whom dalbavancin was introduced beyond 25 days received an elevated number of treatments preceding dalbavancin, considered salvage therapy, and had a prolonged hospitalization (up to 311 days). These patients are presented in the general results and the univariate analysis but were excluded from the comparison with the national database, as they were hardly comparable. For that reason, we chose to present the budget impact according to 1) the 3 main indications (BJIs, IE and ABSSSIs), 2) the number of treatments prior to dalbavancin initiation (1 or 2 or any number), and 3) the LoS before initiating dalbavancin (≤7 days, ≤11 days or ≤25 days), taking into account the catheter type and the severity of the patient’s condition.

LoS was always shorter when dalbavancin was used (Figure 1), up to a reduction of 13 days. A notable exception was when dalbavancin was introduced within 25 days for ABSSSIs, regardless of the presence or type of catheter, or for moderately severe patients, which only concerned 6 and 3 patients, respectively.

**Figure 1:**
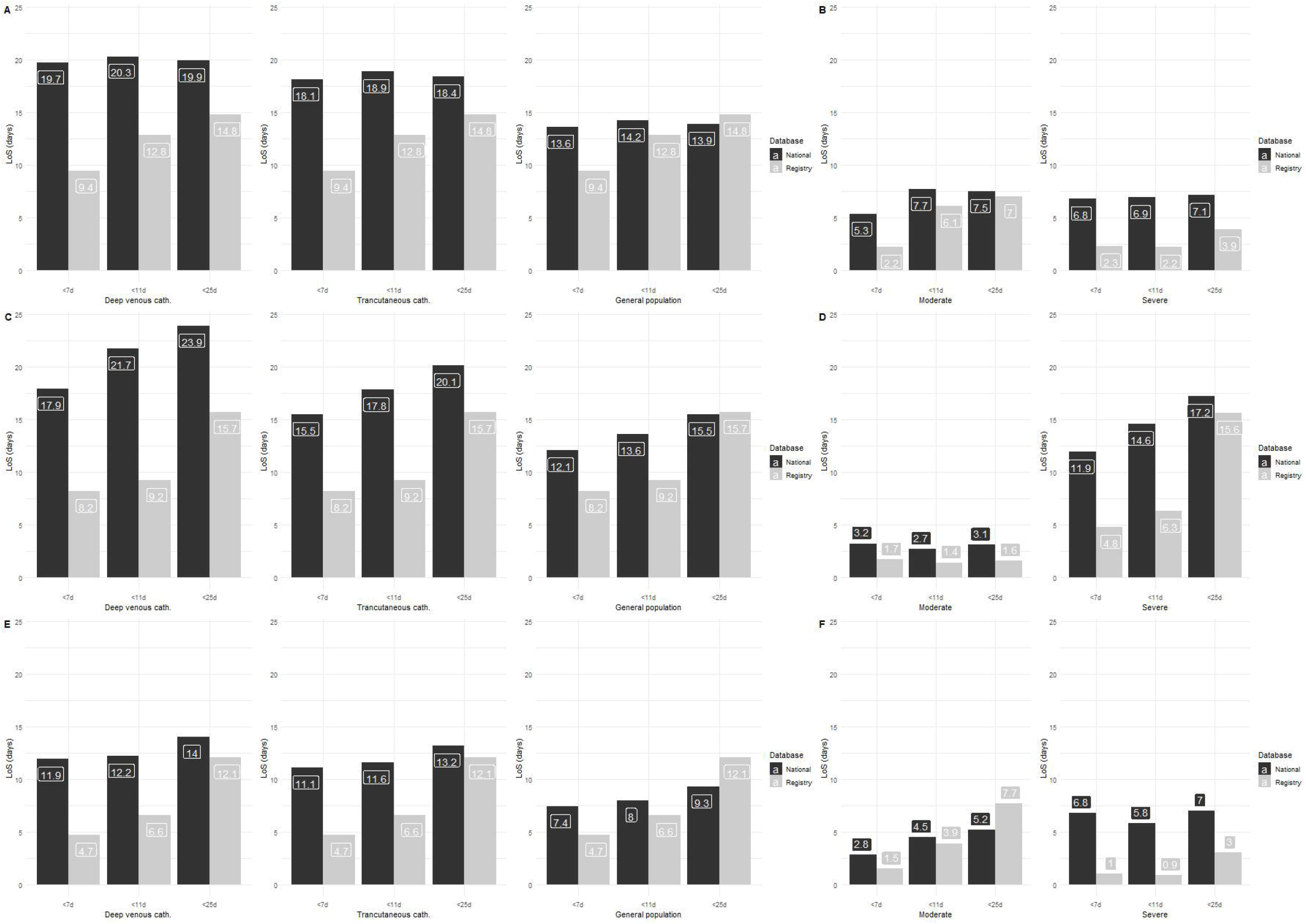
Comparison between the French registry of dalbavancin and the national database of the length of stay according to infection site (A-B: Bone joint and infections, C-D: Infectious endocarditis, E-F: Acute bacterial skin and skin structure infections), catheter type (A, C, E) and severity (B, D, F).

Revenue related to the DRG and additional costs related to the use of dalbavancin instead of vancomycin or daptomycin are detailed in Supplementary Figure 2. Comparison to patients treated with daptomycin was more favourable than with vancomycin treatment.

Dalbavancin generated almost always revenues (up to 3854€) (Supplementary Figure 2), and the resulting budget impact was heterogeneous, from a savings of 2257€ to additional costs of 2227€ (Figures 2 and 3). Early use was associated with savings (or lesser costs). Patients with deep venous catheters (Figure 2) and those with the most severe disease (Figure 3) would benefit the most from dalbavancin. Compared to patients with deep venous catheters, dalbavancin initiated within 7 days would generate savings (or neutral costs) (from 1343€ to -75€). Dalbavancin treatment for the most severe patients was associated with savings when compared to daptomycin (up to 2257€) and vancomycin (up to 2002 €); the notable exception was late use in IE.

**Figure 2:**
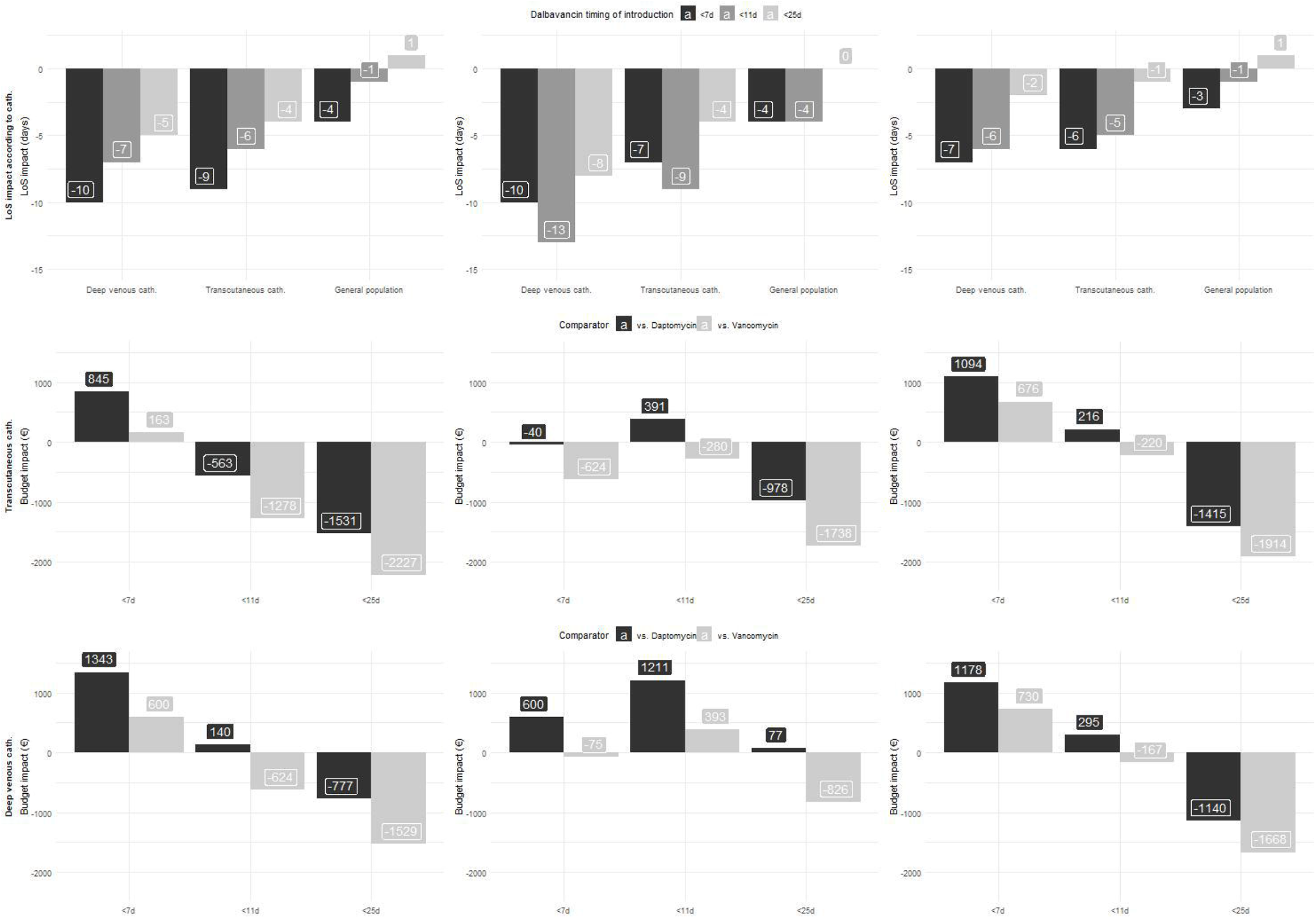
Impact on the length of stay (first row) and budget (second and third row) of dalbavancin according to the timing of introduction, the site of infection (left column: BJI, central column: IE, right column: ABSSSI) and the catheter type (second row: transcutaneous cath., third row: deep venous cath.).

**Figure 3:**
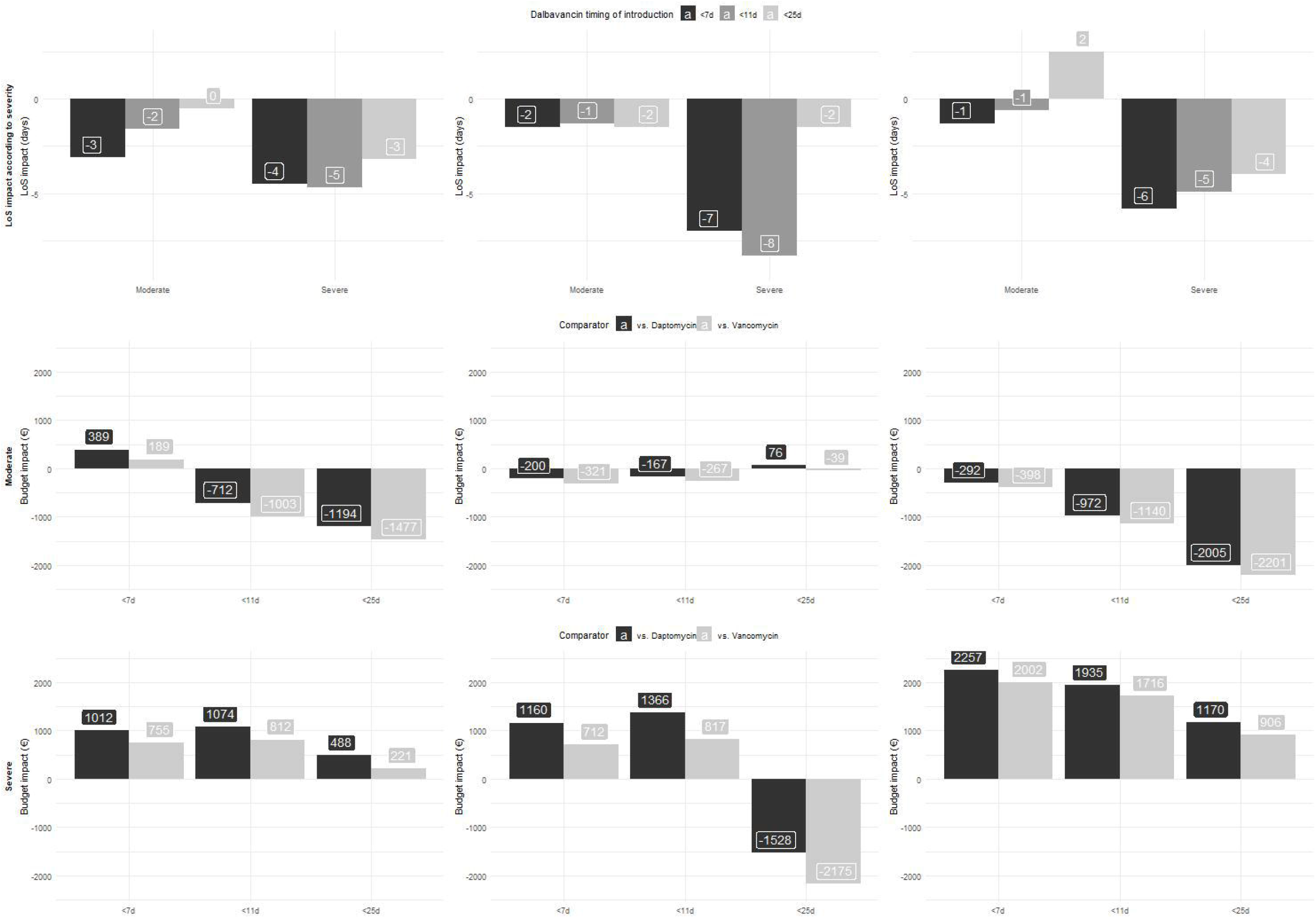
Impact on the length of stay (first row) and budget (second and third row) of dalbavancin according to the timing of introduction, the site of infection (left column: BJI, central column: IE, right column: ABSSSI) and the patient severity (second row: moderate, third row: severe).

A calculator is provided in the supplementary material as a tool to estimate the impact of dalbavancin use.

## Discussion

Dalbavancin use was almost always associated with a sizable reduction in the LoS. Consequently, the resulting budget impact produced savings or extra costs depending on the chosen scenario. To our knowledge, this is the largest study assessing the impacts of dalbavancin on LoS and budgets for multiple indications in real-life conditions.

Since the demonstration of efficacy on ABSSSIs and excellent tolerance with 1000 mg and 500 mg administered a week apart [7], a recent study confirmed comparable efficacy and increased patient satisfaction with 1500 mg given once [21]. ABSSSIs concern a significant proportion of patients, and the prolonged half-life of dalbavancin makes early discharge of the patients possible. In fact, the economic burden of ABSSSIs and the benefit of early discharge in this context have been largely confirmed [22]. Consequently, two modelling studies showed the benefit of dalbavancin on the LoS [12,23] and the economic impact [12] in a European context. In accordance with our results, dalbavancin use was not associated with any additional cost, as the incremental cost of dalbavancin was offset by the decrease in requested resources for its use [12]. Additionally, in accordance with the economic model developed for Germany showing, dalbavancin potential to create an average savings of 2964 € for MRSA ABSSSIs and BJIs [14]. Similarly, a meta-analysis showed that dalbavancin could save third-party payers $1442 to $4803 per complicated skin and soft-tissue infection (cSSTI) [24]. Moreover, potential health risks associated with prolonged hospitalization should be taken into account, as well as the improvement in the quality of life related to early discharge. Beyond the hospital perspective, a small study also suggested decreased direct and indirect costs for self-pay (i.e., uninsured, usually with a low income) patients with ABSSSIs who were switched to dalbavancin after discharge [18].

Although dalbavancin use is supposed to be dedicated to ABSSSIs, it is widely used off-label [25] as a preferred antibiotic due to its efficacy, good tissue penetration and excellent activity against streptococci and staphylococci as well as its long half-life. Our results are also in accordance with an RCT showing a significant and considerable reduction of more than 2 weeks in the LoS for patients treated with dalbavancin for BJIs (15.8 days in the dalbavancin group vs. 33.3 days in the standard of care group (*P* < .001)) [6]. Unsurprisingly, more than half of the patients from our cohort were indeed treated for BJI. This is similar to the study by Bouza et al [16], where dalbavancin was the third antibiotic used, except for two patients for whom it was a second-line treatment and where the standard of care was daptomycin. It should be noted that no patient treated with dalbavancin under compassionate use was included in this study, which has been the preferred use of dalbavancin for a long time. Consequently, and similar to our study, dalbavancin provided an overall cost reduction of 3064 € when used as a second- or third-line treatment. However, the estimation for cost reduction was done by comparison to the theoretical cost of daptomycin treatment for an inpatient based on expert opinion, without comparison to real patients. Another real-life study of dalbavancin use in a US context suggested a mean cost savings of US$40414 per patient [17]. Again, it was compared to a theoretically expected LoS for standard of care. A common reason for choosing dalbavancin was concerns for the PICC line, whether because of drug addiction or another relative contraindication to the PICC line. Nair et al also reported that the cost of the drug was often offset by an earlier discharge made possible by dalbavancin. Finally, a multicentre retrospective study from the USA also showed a reduction in the LoS compared to the usual duration and suggested potential savings but without formal cost analysis [26]. Dalbavancin was then used as a bridge with its usual comparator after a median duration of 13.5 days. In addition to the reduction in LoS, Marcellusi et al also highlighted the reduction in PICC-related adverse events, as PICCs are not necessary for dalbavancin administration, and drug adverse events compared to vancomycin. These results are important from a healthcare insurance perspective due to the economic impact. However, many studies also highlighted the benefit of dalbavancin use in patient quality of life and time savings for healthcare providers, although not formally assessed yet [16,21,27].

By making possible an earlier discharge, the maximum benefit of dalbavancin is obtained with early use (first- or second-line treatment, <7 to <11 days after hospitalization). Implementing dalbavancin in short delays advocates for documented use. Therefore, we suggest that dalbavancin should be recommended as an early documented treatment rather than for compassionate use after failures of other treatments, as many “new” antibiotics are often used.

Another noticeable result is the greatest benefit of dalbavancin use among patients who should have requested a deep venous catheter. Indeed, dalbavancin makes possible the discharge of patients without waiting for the pose of a catheter, which is often the reason for a late discharge of patients. Transcutaneous catheters being easier (and faster) to get than deep venous catheters, dalbavancin provides a more modest benefit. The greater benefit of using dalbavancin among patients with the most severe disease could be related to a better tolerance or efficacy when compared to vancomycin or daptomycin, but we cannot exclude a recruitment bias wherein patients would have been slightly less severe when receiving dalbavancin instead of vancomycin or daptomycin.

Our results also highlight the evolution in dalbavancin use. Bouza illustrated predominantly off-label use in 2016-2017, with only 21.7% of 69 patients treated for ABSSSIs, and dalbavancin was initiated after a median (IQR) of 2 (1-3) antibiotics and 18 (9.8-50.5) days [16]. Dinh et al reported dalbavancin use in 17.3% of patients with ABSSSIs in 2017-2018 in France with a mean (SD) of 2.3 (1.2) prior antibiotics for a median (min-max) of 22.5 (14.3-39.8) days before dalbavancin. Our cohort, gathered in 2019, was very similar to that of Dinh et al except for a lower number of prior antibiotic treatments, with a mean (SD) of 1.6 (1.3) (p<0.001) and a shorter delay of 11.0 (4.0-24.0) days (p<0.001) (Supplementary Table 3). This illustrates the shift towards a more precocious use of dalbavancin.

In the framework of a microcosting approach, similar efficacy between dalbavancin and its relevant comparators, vancomycin and daptomycin, is assumed, and the focus is on the economic impact. Noninferior efficacy has been shown whether as a first-line treatment or as “bridge use”, which could be an early switch or a late switch from various treatments. This could be regarded as a limitation, but the aim of this study was to perform a budget impact analysis based on real-life patients with a microcosting approach, hence the assumption of similar efficacy. Moreover, there are currently not enough data available to stratify precisely according to infection sites and types and pathogens. Finally, there is nonetheless a sizable amount of available scientific literature indicating that dalbavancin is at least noninferior to vancomycin and daptomycin. However, further studies are necessary to identify optimal situations for dalbavancin use and, more importantly, suboptimal situations if any.

We chose to be conservative in our choices to avoid favouring dalbavancin. Consequently, we excluded outpatients, although a significant number of patients were exclusively managed as outpatients, highlighting the benefit of dalbavancin in limiting unnecessary hospitalization days. Specific costs associated with vancomycin, voluntarily excluded as they may vary, would nonetheless decrease the potential extra cost related to dalbavancin use. Similarly, daptomycin costs were calculated for a low dosage of 8 mg/kg/d, while the current trend is to use 10 mg/kg/d. Therefore, we are confident that we did not overestimate the benefit of using dalbavancin but rather provided a minimal expected estimate. Our results also reflect the current mode of financing dalbavancin because of a hospital perspective and an exclusive “intra-DRG” dalbavancin financial model. A probable evolution of financing dalbavancin in its OPAT use (2nd infusion or more) may increase its cost-effectiveness for hospitals. Therefore, a French national payer perspective is needed.

Antimicrobial stewardship has aimed to reduce antibiotic selection pressure by shortening hospital stay to limit nosocomial infections. Therefore, dalbavancin, whether as a first-line treatment or as a “bridge therapy” after vancomycin or daptomycin, should now be regarded as a relevant option for antimicrobial stewardship. This raises the necessity to identify patients for whom dalbavancin use should be anticipated in advance. It could be dedicated to patients for whom long parenteral antibiotic therapy is indicated, such as for those with orthopaedic prosthesis infections or with IE when there is documented microbiological evidence. However, within a more conceptual framework, predictors for the need for a long half-life treatment should be sought, as already published for ABSSSIs [28]. Unsurprisingly, such predictors are in accordance with the literature (IV drug abuse) and our results (high number of comorbidities, severe patients, etc.). We revised the decision tree proposed for ABSSSIs [28] based on these predictors to adapt to the current trends in dalbavancin use and suggested a strategy to help clinicians identify situations where dalbavancin would be useful and potentially cost-effective (Figure 4).

**Figure 4:**
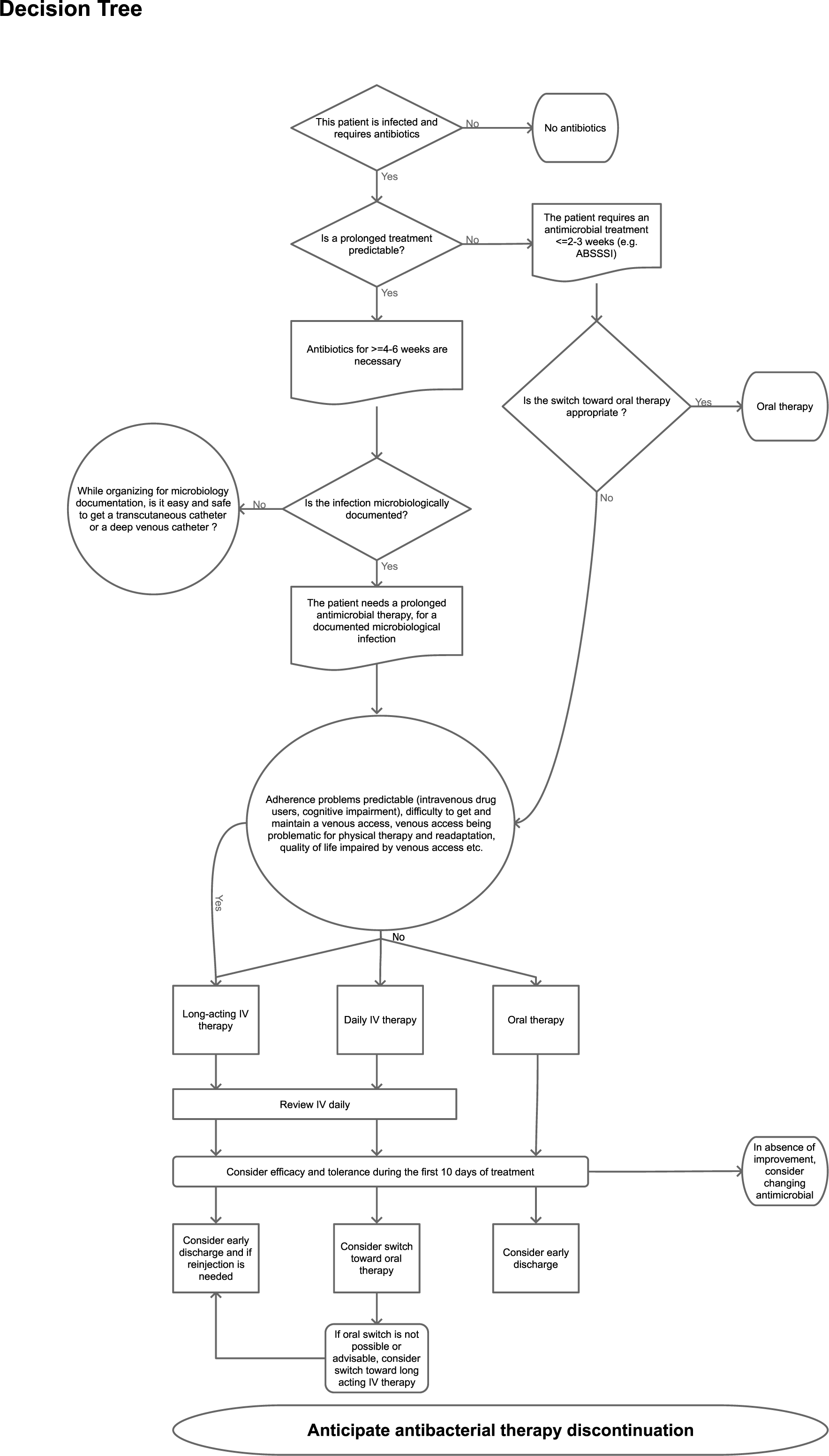
Decision tree to identify situations where long half-life antibiotics such as dalbavancin would be beneficial (derived from Nathwani et al).

The optimal dosing strategy has yet to be defined. A recent systematic review supports either an initial load of 1000 mg followed by a weekly dose of 500 mg or two 1500 mg doses administered a week apart, both being validated by PK/PD studies [11], while 1500 mg once for ABSSSI is also validated [Dunne,HA]. Regarding cost-effectiveness, we suggest the first regimen (1000 mg+500 mg or 1500 mg once) for infections requiring a treatment duration of less than 2-3 weeks and the second regimen (1500+1500) when a treatment of 4-6 weeks is required. More studies are necessary to validate this proposal, but Austrian expert-based OPAT guidelines propose a single shot of 1500 mg of dalbavancin at days 1 and 8 for osteomyelitis and prosthetic infections, regarded as sufficient for an 8-week therapy. For IE, they also recommend 1500 mg at day 1, followed by 1000 mg at day 15, possibly repeated every 15^th^ day.

## Conclusion

Our study confirms that dalbavancin is associated with an earlier discharge of patients, which can offset its costs and produce savings. Moreover, we showed that the benefit of using dalbavancin is always greatest with early use (ideally before the 7th or 11th day of hospitalization) and as a first- or second-line treatment. We therefore suggest that dalbavancin use should be anticipated instead of being dedicated to compassionate use after multiple lines of treatment.

## Data Availability

Data were all public

## Acknowledgements

The authors are grateful to American Journal Experts for editing the manuscript.

## Transparency declarations

The analysis of the UNIHA cohort was conducted by PHE and was funded by Correvio. A.V. and M.B. are employed by PHE. All other authors: none to declare.

## Supplementary material

**Supplementary Figure 1: Establishments participating in the UNIHA 2019 cohort**.

**Supplementary Figure 2: Cost and revenue provided by the use of dalbavancin instead of daptomycin or vancomycin according to infection site (A-B: Bone joint and infections, C-D: Infectious endocarditis, E-F: Acute bacterial skin and skin structure infections), catheter type (A, C, E) and severity (B, D, F)**.

## Dalbavancin Budget Impact Tool

https://github.com/darkdoudou/DalbavancinBudgetImpact/blob/c819519f6f0cd0738966a7cb4a2cb4d80b72bdbd/DalbavancinBudgetImpactTool.xlsx

**Supplementary Table 1:** Comorbidities, pathogens and number of lines of treatment before dalbavancin. The maximum was 12500 for bone and joint infections.

|  | N | Diabetes | Obesity | IVDU | Cancer | Cirrhosis | Poly arthritis | Immuno depression | HTA | Other | MSSA | MRSA | Coag. neg. Staph | MRES | Streptococcus spp. | E. faecium | E. faecalis | Coryne bacterium | Autre | Non identifié |
| --- | --- | --- | --- | --- | --- | --- | --- | --- | --- | --- | --- | --- | --- | --- | --- | --- | --- | --- | --- | --- |
| Bloodstream infections | 10 (6%) | 30% | - | 20% | 60% | 10% | - | 20% | 50% | 60% | 30% | 50% | - | 10% | 10% | - | - | - | - | - |
| Infective endocarditis | 29 (19%) | 10% | 3% | 17% | 14% | 7% | - | 3% | 31% | 38% | 31% | 14% | 17% | 14% | 3% | 7% | 21% | - | 10% | 3% |
| Bone and joint infections | 86 (56%) | 17% | 7% | 2% | 8% | 2% | 14% | 3% | 38% | 50% | 23% | 14% | 9% | 40% | 8% | 5% | 10% | 15% | 15% | 1% |
| Soft tissue infections | 10 (6%) | 20% | 20% | - | 10% | 10% | - | 10% | 30% | 60% | 10% | 10% | - | 10% | 10% | - | - | - | 30% | 40% |
| Catheter-related bloodstream infections | 9 (6%) | - | - | - | 44% | - | 11% | 22% | 33% | 22% | 11% | - | 11% | 78% | - | - | - | - | - | - |
| Endovascular infections | 2 (1%) | - | - | - | - | - | - | - | - | 50% | - | - | - | - | 100% | - | - | - | - | - |
| Others | 6 (4%) | - | 17% | - | 17 | - | - | 17% | 67% | 83% | - | - | 17% | 67% | 0% | 17% | 17% | - | 33% | - |
| Missing | 2 (1%) | 50% | 50 | 50% | - | - | - | - | 50% | 100% | 50% | 50% | - | - | 50% | - | 50% | - | 50% | - |
| Total | 154 | 16% | 7% | 6% | 15% | 4% | 8% | 6% | 38% | 49% | 23% | 15% | 10% | 33% | 8% | 5% | 11% | 8% | 14% | 4% |

**Supplementary Table 2:** Five antimicrobials represented 77% of the preliminary antimicrobials used alone or in combination against gram-positive cocci before dalbavancin was used in the second position. The same five antimicrobials represented 58% of the preliminary antimicrobials used alone or in combination against gram-positive cocci before dalbavancin was used in the third position.

|  | N | L1 | L2 | Post DAP | Post VAN | Post LZD | Post AMX | Post TEC | L3 | Post DAP | Post VAN | Post LZD | Post AMX | Post TEC |
| --- | --- | --- | --- | --- | --- | --- | --- | --- | --- | --- | --- | --- | --- | --- |
| Infective endocarditis | 29 | 1 | 10 | 1 | 1 | 1 | 3 | 0 | 13 | 7 | 1 | 3 | 5 | 1 |
| Bone and joint infections | 86 | 12 | 39 | 13 | 3 | 9 | 5 | 1 | 23 | 9 | 3 | 6 | 3 | 2 |
| Soft tissue infections | 10 | 0 | 7 | 1 | 1 | 0 | 3 | 0 | 3 | 0 | 0 | 3 | 2 | 0 |
| Total | 125 | 13<br>(10%) | 56<br>(45%) | 15<br>(27% out of L2) | 5<br>(9% L2) | 10<br>(18% L2) | 12<br>(21% L2) | 1<br>(2% L2) | 39<br>(31%) | 16<br>(21%) | 4<br>(5%) | 12<br>(15%) | 10<br>(13%) | 3<br>(4%) |

**Supplementary Table 3:**
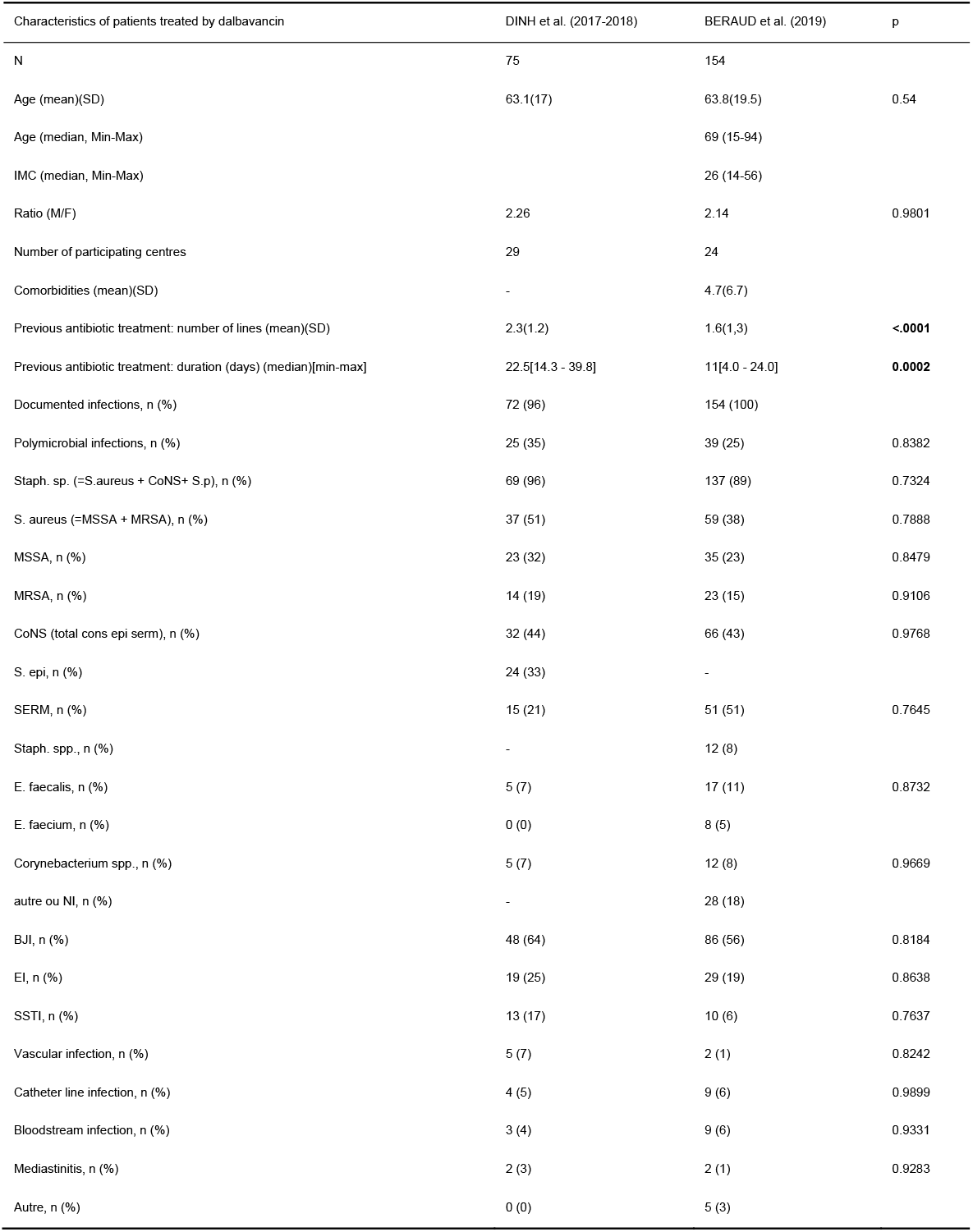
Comparison with Dinh et al.

