## Supplementary figures and images for "Dalbavancin in real life: Economic impact of prescription timing in French hospitals"

### Supplementary Figure 1

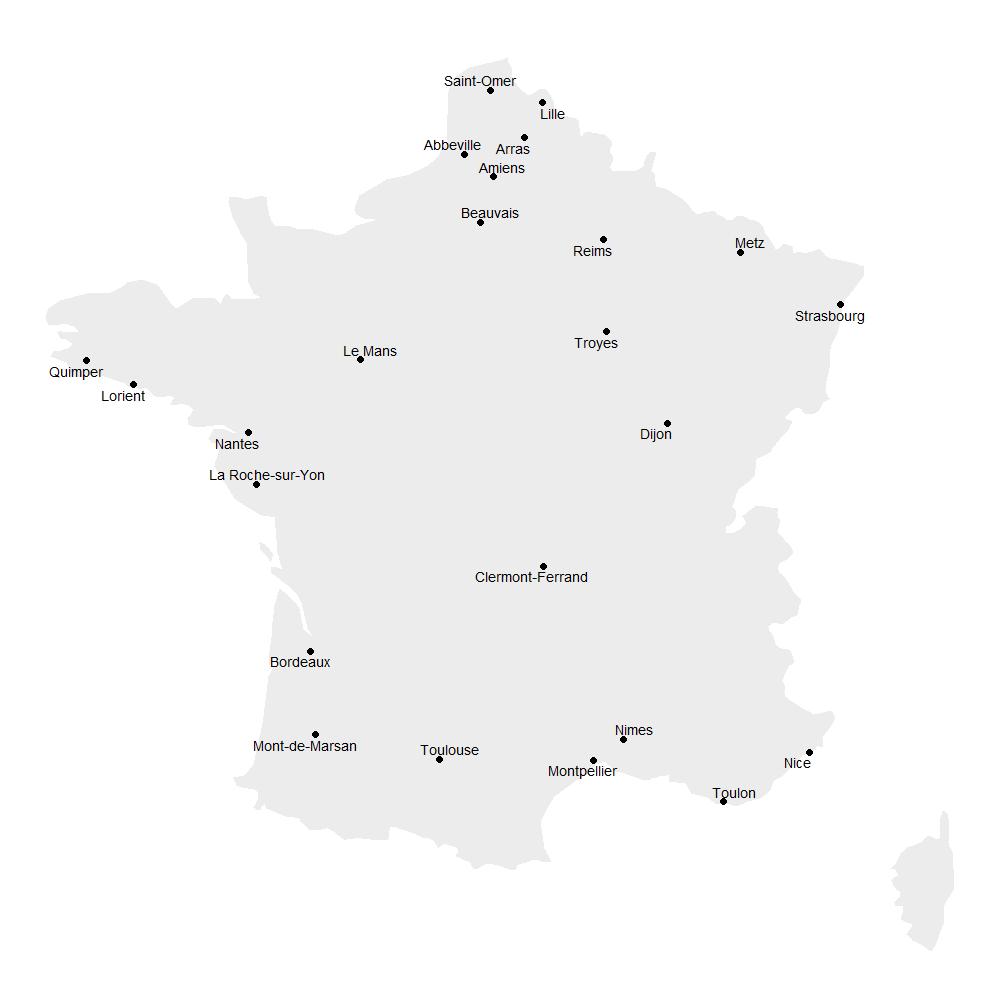

### Supplementary Figure 2

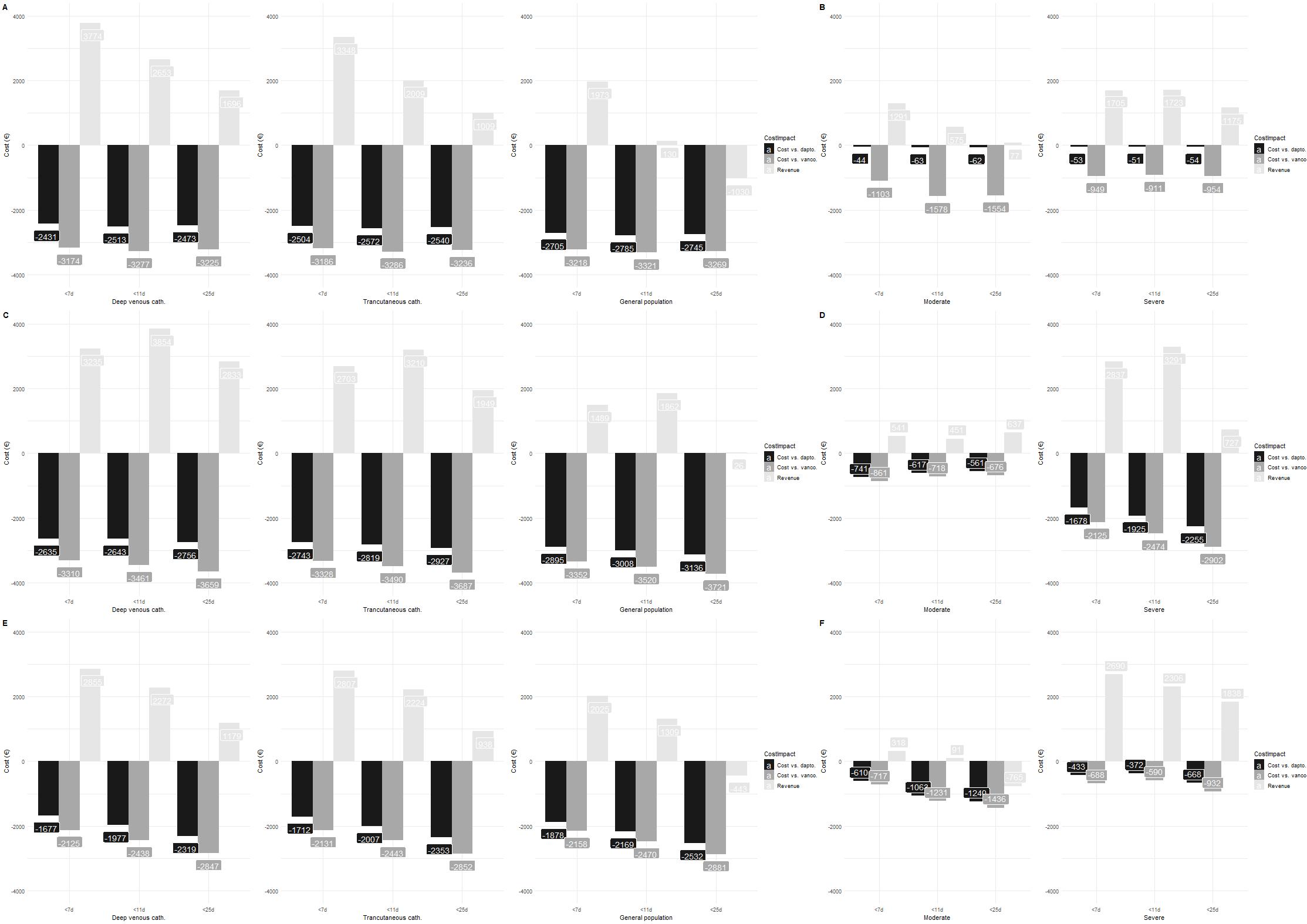
